# Upper limb rehabilitation in facioscapulohumeral dystrophy (FSHD): a patients’ perspective

**DOI:** 10.1101/2020.05.11.20097022

**Authors:** R Kulshrestha, A Faux-Nightingale, N Emery, T. Willis, F Philp

**Author notes:** Corresponding author: Philp F.

## Abstract

**Purpose:** The study aims to identify exercise programmes used by Facioscapulohumeral dystrophy (FSHD) patients in the community, along with barriers and perceptions.

**Methods:** A web based survey, distributed to patients on the UK FSHD registry, and focus groups were conducted. Thematic analysis was conducted on answers to survey questions supported by focus group notes, from seven FSHD patients.

**Results:** A response rate of 43.6% was achieved with 232 out of 532 patients completing the survey. Only 44.4% engaged with exercises targeting the upper body. The themes from the data were: 1) Understanding of disease mechanism shaping exercise choice 2) Lack of understanding about the condition and how exercise interacts with it 3) Support from professionals 4) Barriers to exercise and 5) Thoughts about future research.

**Conclusion:** Exercise selection was variable amongst FSHD patients. Lack of information, pain, fatigue, availability and access to facilities, cost and time were identified as barriers to exercise. Participants (92.2%) agreed additional research into upper limb exercises is needed and felt a 3-month arm cycling intervention with monthly clinical visits and MRI imaging would be appropriate. Further research is needed to develop evidence based exercise interventions and guidance for upper limb exercise prescription in FSHD.

## Main text

### Introduction

Facioscapulohumeral dystrophy (FSHD) is one of the most common inherited muscular dystrophies with an estimated prevalence in general population of 1:21,000 ^[17] [6]^ affecting approximately 2400 people in the UK ^[16]^. It is widely stated as the third most common genetic skeletal muscle disease ^[10]^, after Duchene muscular dystrophy and myotonic dystrophy. FSHD affects the upper extremities and torso, impacting negatively on muscle mass, shoulder mobility and functional ability during tasks. This is most noticeable in functional tasks that require arm elevation above shoulder height ^[5]^, however, functional ability at any level can be affected ^[4]^. Activities of daily living, such as self-care e.g. combing and washing hair, reaching for objects, grasping and lifting are impaired, affecting participation and recreational activities ^[4]^. Muscle weakness of shoulder muscles can also cause instability at the shoulder manifesting as downward translation of the humeral head with respect to the glenoid (sulcus), subluxation and recurrent dislocation which may contribute to the development of shoulder pain. Secondary features of chronic pain and fatigue also have a negative impact on quality of life ^[3, 5]^.

In patients with FSHD, evidence of the efficacy upper limb rehabilitation is limited and does not provide appropriate guidelines for clinical implementation. Exercise prescription for rehabilitation in FSHD patient’s is difficult given the lack of appropriate guidelines, heterogeneity in patient symptom presentation and disease progression, and existence of co-morbidities of pain and fatigue which affect functional capacity. Despite the disease primarily affecting the upper body and associated musculature, existing studies have predominantly investigated the effect of exercise in the lower limbs for rehabilitation ^[9] [19]^. Moderate intensity lower limb exercises are reported to be safe ^[8, 11]^ but detrimental effects are possible if exercise intensity is too high ^[8, 21]^. Current practice for rehabilitation of the upper limb in FSHD patients is unknown and there is no evidence to guide rehabilitation of the upper limb for both exercise type and intensity. Our clinical experience suggests that exercise is appropriate for upper-limb rehabilitation FSHD patients and that current practice is variable. The aims of this study are therefore to identify 1) what exercise modalities patients w4ith FSHD are undertaking in the community as a part of their ongoing rehabilitation, 2) patient’s perceptions of upper limb exercises and any barriers to engagement or compliance and 3) what future research projects would gain the support of patients with FSHD and would be manageable for them.

### Materials and methods

In order to meet the first aim of the study, a web based survey was performed (Appendix 1) The survey was distributed electronically to all patients registered on the UK disease specific FSHD registry ^[12]^. In order to meet the second aim of the study, a focus group was carried out in patients affected by FSHD. Participants were also recruited through the FSHD national registry. All responses were included in this analysis. A thematic analysis was carried out on the open-ended questions from the survey. Due to technical failures, the focus group, which contained 7 participants with FHSD, was not recorded and so no analysis was carried out. However, notes produced during the focus group session were used to support the analysis of the survey data. Data for this study was presented according to the standards for reporting qualitative research reporting guidelines ^[13]^ (Supplemental file 3).

## Results

### Results for response rate to questionnaire

In total, 232 patients completed the survey out of 532 invited patients (43.6%). The average response rate of the fixed answer questions was 99.4%, 193 participants (83.2%) answered one open ended question and 124 participants (53.4%) answered the second. The responses to the open-ended questions were mostly short (one word or a single sentence), with a minority of multiple sentence responses.

### Results for responses related to exercise

The survey results identified that 85.8% of participants agreed that shoulder instability affects their daily life although, despite this, only 44.4% reported engaging with exercises that target the upper body. 32.3% of respondents performed exercises which targeted their upper body more than once a week and the remaining 12.1% performed exercises targeting the upper body less than once a week. This was also echoed in the focus group where the participants identified the value of exercise but conceded that it was “something [that they] don’t do”. Of those who do exercise, there was a wide range of activities and exercises described by participants summarised in table 1.

**Table 1:** Summary of exercises reported by patients with FSHD.

|  |  |
| --- | --- |
| **General physical activity – no additional exercises outside of daily activities/occupational requirements** | Regular movement, no specific exercise, general walking, stair walking, daily tasks/everyday activities |
| **Resistance training with load or assistive machines** | Rubber band/resistance band exercises, low impact- low weight exercises, strengthening exercises, weights, weights for arms, shoulder rope pulleys/ shoulder machines, Ab cruncher, Gym workout/ programme, cable multi-gym |
| **Water-based** | Hydrotherapy/pool based exercises, swimming, Aquafit |
| **Cardiovascular training** | Bike riding, gentle rowing, cross trainer, punch bag, arm cycling |
| **Body weight exercises** | T'ai chi Pilates, Plank, Floor exercises, Wall push ups/ push ups |
| **Maintaining range, mobilising or stretching** | Yoga, Qigong, neck exercises, stretching, Slow stretches, door based exercises, rotation |
| **Core exercises** | Pilates, Floor exercises |
| **Therapist assisted** | Physiotherapy, Osteopathy |
| **Other/ non descriptive** | Arm exercises, palm to palm exercises, deep breathing, adapted game based exercises e.g. throwing and catching, wii sports |

Most long-answer responses included some exercises or activities that the patient carried out, with some individuals also describing the positive impact that it has on their condition, for example in this statement about arm cycling:

> “I already do arm cycling at nmc and I can tell the difference with movement and less aches”

There were very few who described negative repercussions from exercise, and where these were present, they were often related to a specific exercise or piece of equipment.

> “I struggle using a cross trainer due to my FSHDM as the arm movements lead to cramps in my trapezius muscle and severe pain and swelling in both collar bones!”

Patients’ experiences suggest that exercise offers a beneficial mechanism to improve flexibility and strength and that a wide range of exercises can be utilised to target different locations in the body, including the upper limbs. Responses vary between patients, suggesting that these exercises help patients differently according to their symptoms.

Following the application of thematic principles, the following themes were constructed from the data

1. Understanding of disease mechanism shaping exercise choice
2. Lack of understanding about the condition and how exercise interacts with it
3. Support from professionals
4. Barriers to exercise
5. Thoughts about future research

### Understanding of disease mechanism shaping exercise choice

Patients’ often accompanied their list of exercises with explanations of how it supports or interacts with their condition, emphasising the knowledge and understanding that they have in this area. This desire to work in order to counter the effects of FSHD, or patient empowerment through acquired knowledge, is common in the survey responses: patients who, having been diagnosed with a degenerative illness, research it to understand what it does and use that knowledge to determine what exercises are beneficial to them. For example, here, where a participant discusses the different exercises that they undertake and the benefits they hope to gain as a result:

> “1. Pliates to improve core muscle strength 2. Yoga to maintain flexibility, and to enable arm cycling while braced on the floor 3. Walking whilst possible”

Similarly, there is a willingness from patients to try new things which might help improve their condition. Many responses in the survey mention exercises or equipment that they have tried, or previous research studies that they have engaged with, with varying levels of success. Patients with FSHD use this accumulated knowledge to assess the potential suitability of exercises or machines. This extends to items that have already been used with problematic results and there seems to be a willingness to try again in case a correctable mistake was made previously or the item has been adjusted in some way:

> “I have tried to use that piece of equipment several times over the last decade, unfortunately each time it has caused severe pain and breathing difficulties each and every time. Having said that it has never taken place under supervision, so I think with the correct set of circumstances it is definitely worth a trial.”

Patients have sought to learn about FSHD to a point where they understand it and how exercises can help moderate the symptoms, this not only helps them physically but also gives them the tools to help themselves in the future. It also allows them an element of control and hope for the future which can impact on their psychological wellbeing.

### Lack of understanding about the condition and how exercise interacts with it

Of the participants who do not engage with targeted upper limb exercise, there are two main groups 1) those who do no specific or additional exercise outside of daily activities/occupational requirements, for example,

> “do no specific exercise, [finding] regular movement during the day […] is of much benefit to the maintenance of my remaining shoulder movement”,

and those who don’t do any exercise or activities for physical benefit. Of the participants who don’t engage with any type of exercise or activity, a notable proportion report not exercising due to confusion surrounding the exercises or a lack of knowledge about their condition. These views were captured by the following comments:

> “Unsure, as have never had any correct exercises suggested.”
>
> “I would definitely benefit from arm exercise but do not know what type would be possible.”

The impact on wellbeing and quality of life is presented in some of the survey responses with patients reflecting on their lack of knowledge and the impact that it could have had on their life had they known about it, for example:

> “Not sure but wished I knew more on what i can and cant do [sic]”
>
> “I haven’t done any exercises at all over the years because I accepted my condition and didn’t realise that exercise would help…”

This concern about the availability and quality of information which might help an individual with FSHD was mirrored in the focus group where participants were concerned that professionals don’t all have an equal understanding of the condition or associated problems. Patients were concerned about this and were worried that this was leading to inconsistent advice and exercise recommendations

The lack of understanding surrounding both the exercises and the condition impacts on the patients physically and psychologically. Physical implications stemming from a lack of understanding prevented some patients from improving their strength and mobility, whilst psychologically, patients are denied control over their wellbeing and support with their life-controlling condition. Patients in the focus group identified that this was a key area that needed to be rectified, reporting that the system needs to be adapted so that professionals explain symptoms and medical information thoroughly and in a way that is accessible to patients. The focus group also agreed that they would benefit from more information about all aspects of the condition, alongside relevant exercises or equipment to support them and sign posts provided for further information or professionals which may help.

### Support from professionals

A number of survey responses mention “exercises recommended by my physiotherapist” and the support that they receive from hospitals. However, a greater number mention advice and recommendations that they receive from professionals outside of the conventional hospital or national health settings. Patients describe a number of different information sources, mainly alternate professions, which they use to guide their exercises, for example, Pilates instructors, gym instructors, personal trainers, and osteopaths. This is often coupled with an explanation of how the action benefits their strength or mobility, for example:

> “Working with a level 4 gym instructor, we have been targeting exercises to increase movement of arms to raise to shoulder level and above. We have been trying to isolate specific muscle groups and working to strengthen these.”

The additional support that patients receive from external professionals often features in place of any physiotherapy-based recommendations. This is possibly linked to perceived lack of knowledge from medical professionals. Patients reported feeling that the exercises or advice received was lacking or varied between clinicians in conventional hospital or healthcare settings and so sought to get their support from other people with perceived similar training.

### Barriers to exercise

Lack of knowledge was not the only element that contributed to patients not exercising. In both the survey and focus group, a number of factors or barriers which influence patients’ propensity to exercise were identified, namely loss of mobility, reduced functional ability, pain and fatigue. Functional limitations associated with the disease condition of FSHD were the most common barrier raised. Many participants in both the survey and focus group noted that their condition affected their travelling ability, therefore, limiting their ability to engage with exercise regimes or research studies, saying for example

> “I am unable to travel now, and would be unable to take part, other than to give my comments as above.”

These limitations are frequently identified in the survey responses, with many patients offering stipulations in their assessment of exercises, such as “doesn’t involve”, “limited in what I’m able to do” or “progression of disease made it impossible”, “used to do this until”. The focus group echoed these comments and also mentioned problems with increased fatigue levels that can be induced by exercise and which can affect daily tasks or activities during the following hours or days. Whilst this barrier is not unexpected, given the nature of the disease and its progression, it highlights the difficulties associated with exercising in FSHD.

Pain from exercise or problems with exercises was also a significant barrier discussed in the survey responses. A number of patients described situations where particular exercises or machines have “caused severe pain and breathing difficulties each and every time”, some of which resulting in “[having] to refrain from even trying for the last 4 months.”. Although problematic, this was not always a complete barrier, as many of the responses reported continuing to exercise areas of their body which were pain-free.

In addition to the condition based barriers, both the survey responses and focus groups mentioned availability and access to facilities, whether through cost, location or time, as a significant barrier contributing to lack of exercise. Although many of these barriers raised cannot be easily overcome, discussion within the focus group offered some suggestions of ways to improve exercise engagement. These included using adjustable height tables to reduce discomfort while training, the distribution of an endorsed regime which would ensure consistent, beneficial exercise sessions, and the inclusion of some type of distraction, for example TV or music, to accompany the exercise. These recommendations notably focused on improving the process of exercising itself, rather than overcoming barriers which might prevent exercise, and could be used in a variety of exercise environments, whether at home or in the gym.

### Thoughts about future research

The vast majority of participants (92.2%) agreed that there was need for additional research into upper limb exercises for patients affected by FSHD. Most patients (93.5%) were happy for studies to take place with clinic visits occurring on a monthly basis, although the majority said that they would like travelling expenses to be covered. With regards to the study intervention, 56.9% of patients agreed that arm cycling would be a feasible exercise for their condition and 38.8% were not sure. A three-month arm cycling programme was considered a manageable length of time for the intervention by 73.7% of participants and 74.6% of participants reported that they would be willing to undertake an MRI scan as part of a study. The focus group discussion also supported this, with patients agreeing that they would be willing to travel and take part in a study which lasted at least three months, particularly if they could provide their employer with a letter enabling time off to participate. Further discussion in the focus group also suggested that exercises should be gradually made more difficult and that positive reinforcement should be built into the programme, possibly through the use of a goal-based system, to make it more engaging.

## Discussion

The aims of this study were to identify 1) what exercise modalities patients with FSHD in the community are undertaking as a part of their ongoing rehabilitation, 2) patient’s perceptions of upper limb exercises and any barriers to engagement or compliance and 3) what future research projects would gain the support of patients with FSHD and would be manageable for them. We achieved the aims of our study, having identified the range of exercise modalities undertaken by patients with FSHD and proportion of patients who use exercise as a part of their upper limb rehabilitation. We were also able to identify patient’s perceptions of upper limb exercises and associated barriers for engagement and compliance which can be used for informing management and future research.

Within our study it was identified that more than 50% of the respondents were not engaging in upper-limb exercises, despite more than 80% of patients reporting instability which affects their activities of daily living. Of the patients who exercised, a broad range of exercise modalities were identified. Whilst variability in exercise selection may be reflective of the heterogeneity within this patient group, our study suggests that the variability may reflect the lack of understanding regarding FSHD and how exercise interacts with it. Exercise interventions should aim to addresses the main mechanisms associated with the disease pathophysiology or subsequent symptoms and consider the limitations associated with the condition to allow it to be delivered safely and effectively. It is important to note that in some cases, patient’s recalled transient complications associated with exercise. It is recognised that there is insufficient evidence about the effectiveness of all exercise programmes exercise in FSHD and detrimental effects can be experienced depending on the exercise protocol selected ^[8, 21]^. Whilst some exercise programmes have demonstrated effectiveness, for example, increased cardiovascular fitness in FSHD ^[2, 14]^ and other neuromuscular disorders ^[15, 18]^ safe and effective parameters for other exercise protocols are yet to be established. Insufficient levels of physical activity are also associated with increased risk of co-morbidities ^[1]^. It is therefore important that when developing or investigating exercise interventions in FSHD, that these have appropriate physiological rationale.

Patient responses indicate that FSHD patients are willing to engage in exercise and our study supports exercise as a feasible intervention for upper limb rehabilitation. The benefits of exercise, both physical and psychological, are recognised in FSHD ^[20]^ and many other chronic conditions ^[7, 21]^. Physical benefits from lower limb exercises are also recognised in FSHD ^[21]^ and it is therefore reasonable to assume that there are similar benefits that may be achieved for the upper limbs in FSHD patients ^[20]^. The inability to engage with exercise was identified as having both physical and psychological implications. Most notably patients were denied control over their wellbeing and support with their life-controlling condition. Whilst arguably some exercise paradigms may have limited therapeutic potential e.g. strength due the pathophysiology and degenerative nature of the condition, there are likely to be additional benefits associated with exercise and it is important these are accurately captured. Given the potential benefits associated with exercise may have therapeutic benefit it is important to recognise barriers which may prohibit engagement and compliance with exercise.

Lack of information about FSHD and exercise is an already established issue, with a lack of appropriate guidelines available for clinical decision making regarding exercise prescription. It was interesting to note that there was a perceived a lack of knowledge and inconsistency in advice from medical professionals reported by patients which was identified as a factor in them seeking support from other people with perceived similar training. It is probable that patients will be looking for confidence in practitioners to help them manage their condition. Given the realistic uncertainty associated with management of FSHD, the perceived lack of knowledge may therefore be reflective of clinicians navigating decision making in domains of true ambiguity. Availability of appropriate information and shared decision making is therefore essential for management of FSHD. This is particularly important given the degenerative and long-term nature of the condition, as patients with FSHD will be reviewed by medical professionals throughout the course of their condition and also be required to self-manage. It is therefore important that FSHD patients and respective medical professionals have access to sufficient information to allow for the condition to be appropriately managed and foster good therapeutic alliance.

Secondary complications associated with the condition, namely loss of mobility, reduced functional ability, pain and fatigue were also identified as barrier to upper limb exercise in FSHD patients. In addition to the condition based barriers, both the survey responses and focus groups mentioned availability and access to facilities, whether through cost, location or time, as a significant barrier contributing to lack of exercise. It is therefore important that when designing exercise interventions for future studies that these factors are taken into consideration.

Further research is therefore needed to develop appropriate, evidence based exercise interventions and guidance for informing exercise prescription for upper limb rehabilitation in FSHD. In absence of pharmacotherapy or gene therapy, upper limb rehabilitation is the fundamental in the ongoing management of FSHD. Overall, participants in the survey and focus group were positive and agreed about the need for future research into exercises for patients affected by FSHD. The results of our study provide support for the use of a 3-month arm cycling intervention, performed at home or in the clinic, as well as the use of MRI and frequency of clinical visits. The barriers and recommendations identified by patients will also be important for designing research studies, as mobility, pain, access, motivation and fatigue were all identified as factors which may negatively impact on research design, recruitment and overall translation into clinical practice.

### Limitations of our study

Within our study we achieved a response rate of 43.6% for the questionnaire. This is less than half of all FSHD patients, however, given that respondents were recruited from an established UK registry, the responses received are likely to be representative of the population. It is recognised that our study could potentially underestimate the number of people who are not undertaking upper limb exercises, as respondents are more likely to be engaged with exercise and therefore have higher levels of functional capacity. Further work may therefore be required to identify exercise interventions for lower functioning patients or methods for engaging FSHD patients not currently doing exercise. Whilst responses from our study provide anecdotal evidence for the use of upper limb exercises in FSHD, we were unable to objective thresholds for informing exercise prescription or avoidance of complications. Due to technical failures, the focus group transcripts were not available. Notes produced during the focus group session were used to support the analysis of the survey data but it is recognised that analysis of the focus group transcriptions, if available, would have allowed for a more in depth analysis of patient perspectives regarding exercise in FSHD and recommendations for further research.

## Data Availability

Data are available on request

**(Supplemental file 1) - ICMJE Form for Disclosure of Potential Conflicts of Interest**

**(Supplemental file 2) - R&D letter in support of publication of study data 160420**

**(Supplemental file 3) - Completed SRQR checklist**

## Acknowledgements

We express our sincere thanks to FSHD registry, UK and Neuromuscular Centre, Winsford, UK.

## Ethical approval statement

Research and Development approval for this work has been received from The Robert Jones and Agnes Hunt Orthopaedic Hospital NHS Foundation Trust. No ethical approval was needed as per the principles set put in NIHR involve (Supplementary file 2)

